# Physiotherapy Educational Intervention for Occipital Positional Plagiocephaly Prevention: a Multicentre Randomised Controlled Trial

**DOI:** 10.64898/2026.09.03.26362040

**Authors:** Viola Fortini, Fabiola Picone, Simona Giustini, Patrizia Fanzaghi, Martina Santoni, Massimiliano Calistri, Riccardo Guarise, Nicola Ferri

**Author notes:** Corresponding Author: Viola Fortini, PT, MSc,Department of Health Professions Azienda USL Toscana Centro 51100, Pistoia, Italy.

## Abstract

**Questions:** What is the effectiveness of a prenatal educational physiotherapy intervention in reducing the prevalence of positional occipital plagiocephaly (POP) and POP-associated disorders in infants at 3 months of age?

**Design:** Two-arm assessor-blind multicentre, randomised controlled trial.

**Participants:** 400 pregnant women

**Intervention:** The experimental group received specific physiotherapy educational session on POP prevention strategies during the regular antenatal education class meeting. The oblique diameter difference index (ODDI) is the primary outcome. Secondary outcomes included musculoskeletal POP-associated conditions.

**Results:** Three hundred twenty-eight newborns completed the study. The ODDI scores showed no statistically significant difference between the groups at 3 months (mean difference −0.28 [95% CI: −0.72 to 0.17; p = 0.218]). The prevalence of POP did not differ between the two groups (21.1% in the intervention group, 22.2% in the control group), which is lower than data reported in the literature (37.8%–46.6%). No statistically significant between-group differences were found in secondary outcomes, except for spinal postural asymmetry (aRR = 0.95; 95% CI: 0.90 to 0.99; p = 0.031). There were no significant differences in the caregiving strategies adopted during the first three months of life.

**Conclusion:** A low POP prevalence was observed, with no statistically significant differences between groups, as both groups adopted preventive strategies. The lack of superiority of the structured education may suggest a shift in parental education. With more health information on social media and the global ease of sharing, parents have relied on digital self-treatment, reducing the intervention’s impact; thus, the face-to-face model for low-risk infants may be outdated.

**Registration number:** NCT07182604 (retrospective). The protocol was prospectively registered at the Meyer Children’s Hospital IRCCS (January 20, 2020).

## Introduction

### Background and rationale

Positional occipital plagiocephaly (POP) is a skull morphological abnormality that occurs in the absence of early synostosis of the cranial sutures and is caused by external forces acting on the skull, which is highly malleable in newborns.^1^ Children with POP have a skull deformity with oblique craniofacial modelling, characterised by flattening of the posterior occipital region and a same-side anterior protrusion. In rare cases, there is also bilateral POP or brachycephaly, characterised by bilateral occipital flattening of the skull, which is more common in Asian populations due to their phenotypic characteristics.^2^

POP is considered congenital when the head is asymmetrical immediately after birth and requires medical attention if the oblique diameter difference index (ODDI) measured with a craniometer exceeds 112.6%. Indeed, a certain degree of cranial asymmetry at birth is physiological, resulting from mechanical forces exerted during delivery. More often, however, deformities are absent at birth and become apparent in the first 2-4 months of life,^3^ particularly at 7 weeks. In this case, POP is defined as acquired, with peak severity occurring around 4 months of age. Acquired POP with ODDI ≥ 104% requires treatment,^4^ which is mostly represented by physiotherapy,^5–7^ the first-line treatment,^8^ and helmet therapy.^9–12^

The incidence of POP has been estimated at 46.6% at 7-12 weeks of age, and 78.3% of POP cases are considered mild.^13^ By contrast, the prevalence of POP in healthy newborns is 13% at birth, rising to 16% at 6 weeks,^3^ and 37.8% at 3 months,^14^ then decreasing to 3.3% at 2 years.^15^

Since 1992, there has been a significant increase in the diagnosis of POP, with a more than sixfold increase between 1992 and 1993.^16^ This is likely related to the recommendations of the American Academy of Paediatrics (AAP) through the "Back to Sleep" campaign, which advises placing newborns on their backs to reduce the risk of sudden infant death syndrome (SIDS).^17–19^ New hypotheses have recently been proposed, according to which the increase in the incidence of POP seems to be due more to a failure to promote movement in newborns, especially the lack of habit of placing children in the prone position while awake, rather than solely to the supine position.^3,4,20–22^

The main risk factors for POP are divided into maternal, perinatal, and postnatal factors. In summary, male gender, primiparity, fetal malposition, and difficult, prolonged labour are conditions that increase the risk of congenital POP.^4,15^ The risk factors for acquired POP at 7 weeks of age are: male gender, primiparity, preferred sleeping position, the newborn’s head-to-foot position in the crib always in the same direction, the position during bottle feeding always on the same side, the prone position while awake used less than once a day, and a delay in motor skill acquisition.^4^

In addition to being an aesthetic problem that affects the shape of the skull and face, POP can be associated with children’s postural-motor development,^23^ muscular imbalances and postural asymmetries in the neck, such as postural torticollis, or spinal postural asymmetries, or asymmetries in the functional use of motor skills.^24^ Frequently, attention is paid only at a late stage, when the situation is very evident and often associated with other problems, resulting in longer, more expensive physiotherapy treatments with poorer outcomes. Although it is not a serious condition, it is a very common problem in the first months of life and, therefore, very frequent in rehabilitation services for assessment and physiotherapy treatment, resulting in high costs for health care systems. Recently, interest has emerged in the possibility of preventing POP through simple guidelines that can be shared with parents, as also reported in the recent guidelines on the treatment of congenital myogenic torticollis.^25^ Some studies show that preventive and educational interventions with families on the most appropriate ways to care for their children in an appropriate environment, carried out after the birth of the child, effectively reduce the incidence and severity of POP in the first months of life,^26^ and that good nationwide training of healthcare professionals on this subject could help minimise public healthcare costs.^15^

### Objectives

The aim of this study was to assess whether an educational physiotherapy intervention, as part of the birth preparation course, reduces the prevalence of POP in infants at 3 months of age. Secondary objectives were to assess the effectiveness of preventing POP-associated disorders (postural torticollis, neck muscle contracture, spinal and pelvic misalignment, immaturity in anti-gravity axial control) and the quality of the intervention as perceived by caregivers.

## Methods

### Trial design

This is an assessor-blind, multicentre, randomised controlled trial investigating the effectiveness of a physiotherapy educational intervention for preventing occipital positional plagiocephaly, using a superiority framework with parallel groups. This study is reported following the Consolidated Standards of Reporting Trials (CONSORT 2025) checklist.^27^ The trial was approved by the Meyer Children’s Hospital IRCCS Ethics Committee (prot. POP2019,24/09/2019) and registered with the same Hospital (20/01/2020) and on *ClinicalTrials.gov (NCT07182604).* All participants provided written informed consent before participating in the study.

### Trial setting

This multicentre trial was conducted at four hospital centres in Italy: AOU Careggi (Firenze), Infermi Hospital (Rimini), ASL 4 Liguria (Chiavari), and Hospital of Parma (Parma), with the AOU Careggi being the coordinating centre.

### Eligibility criteria

All parents who attended the prenatal classes during the 34th to 36th weeks of gestation were included from January 2023 to October 2023. Parents under 18 years old or with difficulties in Italian language comprehension were excluded. Then, newborns were excluded if presenting at birth with any of the following: prematurity (< 36 weeks), neonatal suffering, congenital POP with ODDI > 112.6%, clavicle fracture, congenital clubfoot, metatarsus varus, or abnormalities of the spine and pelvis (e.g., congenital hip dysplasia, congenital torticollis, cephalohematoma).

### Intervention and comparator

In the intervention group, during regular antenatal education class meetings, information about POP and how to prevent it was shared with parents. A physiotherapist carried out this session through the presentation of a video (Supplementary File 1), the sharing of a POP prevention brochure (Supplementary File 2) which was developed by Italian Group of Study in Pediatric Physiotherapy, followed by practical demonstrations of the strategies using a doll, and then a time of direct discussion between the physiotherapist and parents to clarify any doubts and questions they may have. At the end of the educational talk, participants were given a copy of the brochure. The brochure and the video had previously been approved by a focus group of parents and paediatric physiotherapists.

The control group regularly attended the traditional birth coaching course, thus without receiving any specific information on POP prevention.

All children participating in the study were assessed for eligibility within 48 hours of birth by a physiotherapist who was different from the one who delivered the educational intervention. A predefined form was used to collect data during the birth assessment.

### Outcomes

The primary outcome was the ODDI measured with the calliper *Mimoscraniometer* (Think Pipeline, SLU, Manresa, Spain), which has demonstrated excellent reliability and accuracy.^28–29^ The ODDI is defined as 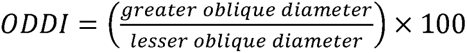. Secondary outcomes included the presence of associated postural torticollis, neck muscle contracture, postural asymmetries of the spine and pelvis, and axial anti-gravity control of the head and trunk. Data were also collected on care practices used during the study period that are relevant to the prevention of POP, in particular infant sleep position, methods of transport (baby carrier, sling, baby bouncer, infant car seat), and positioning pillows. All outcomes were collected at 3 months by an independent physiotherapist. The parents’ perceived quality of the educational intervention was also assessed using a predefined questionnaire (Supplementary File 3) that focused on the usefulness and clarity of the information provided, as well as any difficulties encountered in implementing the prevention program.

The follow-up medical history also investigated any treatments the child had undergone during the research period (e.g., physiotherapy) to address potential confounding factors.

### Sample size

Based on the difference in the prevalence of plagiocephaly observed in a previous pilot study (14,28% in the intervention group and 35,29% in the control group), the required sample size for detecting a significant difference between groups was calculated with an α of 0.05 and a power of 0.85. Given the cluster-randomised design, the sample size was adjusted for within-cluster correlation, resulting in a total required sample of 356 newborns. Adjusting for an estimated dropout rate of 20%, the total sample to be included is 427 newborns.

### Randomisation

An off-site researcher performed cluster randomisation stratified by centre, based on the dates of the prenatal classes.

### Blinding

Physiotherapists who performed the assessments at birth and at 3 months were different from those who performed enrolment or prenatal educational intervention and were unaware of the allocation. Furthermore, although the parents could not be blinded due to the nature of the intervention, it is reasonable to consider the newborn blinded to the allocation.

### Statistical analysis

Baseline characteristics of the sample were summarised by randomisation group. Continuous variables were assessed for normality using the Shapiro-Wilk test and visual inspection of kernel density plots, and were reported as means with standard deviations, or medians with interquartile ranges, as appropriate. Categorical variables were reported as absolute frequencies and percentages.

To appropriately handle missing data and reduce potential attrition bias, we performed multiple imputation using chained equations. Fifty imputed datasets were generated using predictive mean matching for continuous variables and logistic regression for categorical variables. The imputation model included treatment group, hospital centre, baseline ODDI score, birth weight, gestational age, infant sex, and all primary and secondary outcomes. All subsequent pooled analyses were performed according to Rubin’s rules.

The primary continuous outcome (infant ODDI score at follow-up) was analysed using a linear regression model (ANCOVA approach) adjusted for the baseline ODDI score and hospital centre. To account for the cluster-randomised trial design (with clusters defined by the prenatal class groups), cluster-robust standard errors were employed in all regression models to adjust for intra-cluster correlation. Additionally, the intraclass correlation coefficient (ICC) was estimated using a linear mixed-effects model on complete cases to quantify the cluster effect.

For the dichotomised primary outcome (prevalence of plagiocephaly, defined as an ODDI score ≥ 104) and other binary secondary outcomes, modified Poisson regression models with a logarithmic link function and cluster-robust standard errors were used to directly estimate adjusted Relative Risks (aRRs) and 95% Confidence Intervals (CIs). Ordinal secondary outcomes (head and trunk antigravity control) were evaluated using ordinal logistic regression models, similarly adjusting for baseline values, hospital centre, and clustering. Non-parametric sensitivity analyses based on outcome ranking were also performed to ensure the robustness of the primary findings.

All statistical analyses were conducted using Stata software, version 19.5 (Stata Corp, College Station, TX, USA). A two-sided p-value < 0.05 was considered statistically significant.

## Results

Four hundred pregnant women were randomised across the 4 centres; 339 newborns met the eligibility criteria and were enrolled. Eleven participants did not complete the study, resulting in a 3% drop-out rate (Figure 1).

**Figure 1.**
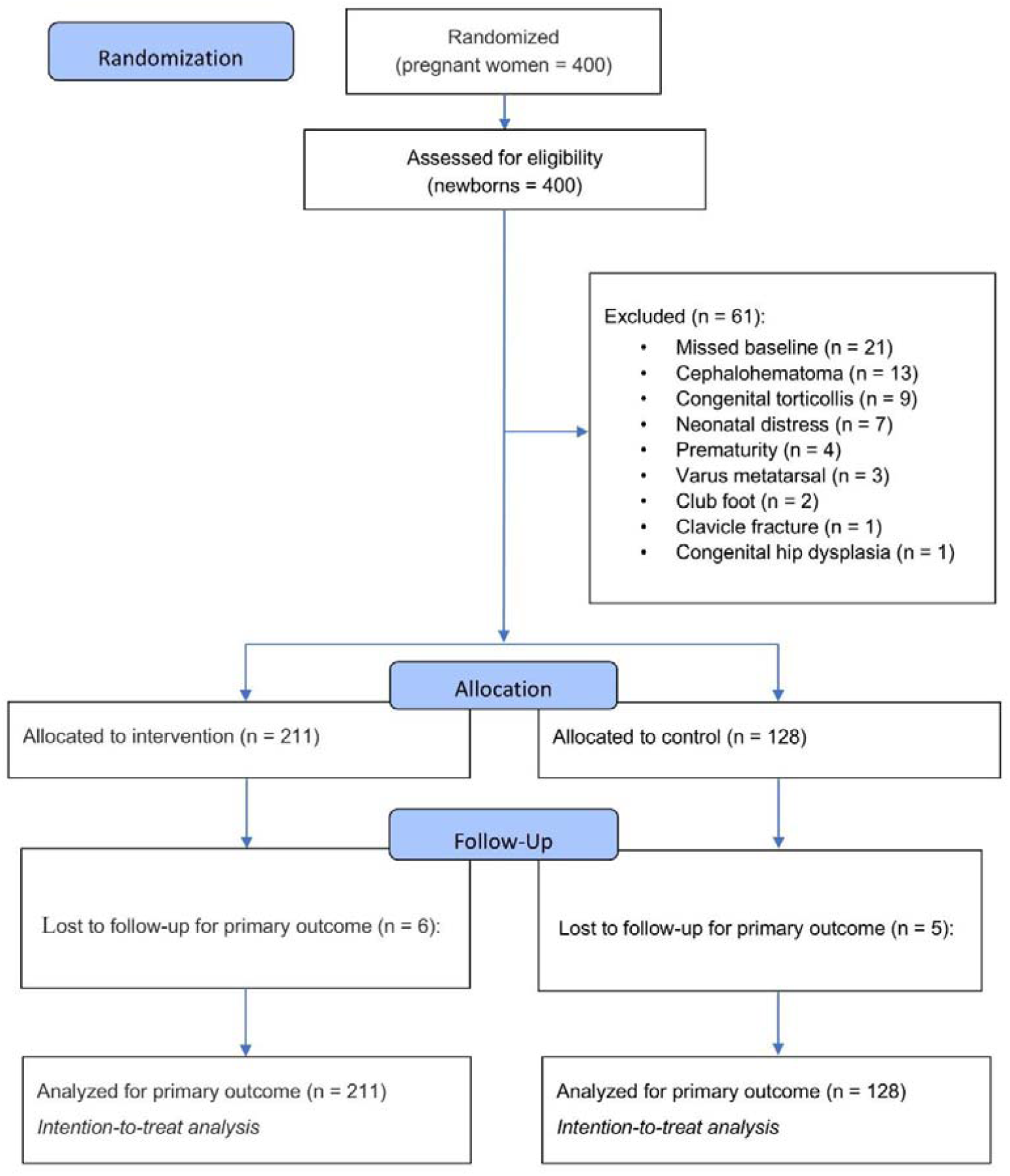
Study flow diagram. The distribution of the sample across centres of both pregnant women and eligible newborns is reported in Figure 2.

**Figure 2.**
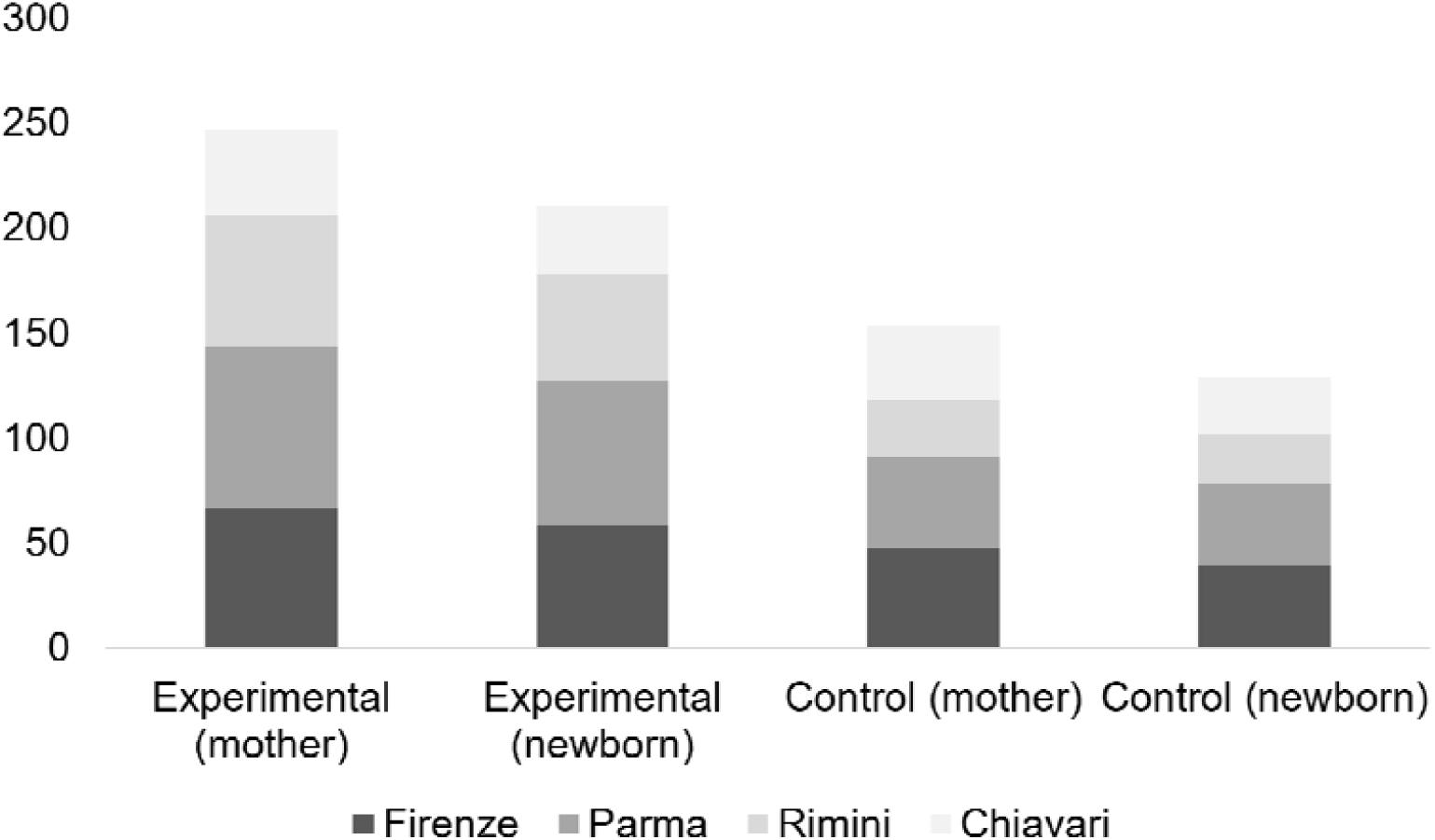
Sample distribution across centres. Pregnant women and newborns in the intervention and control groups were similar in baseline characteristics, as reported in Tables 1 and 2, respectively.

**Table 1.** Pregnant women’s baseline characteristics.

|  | Intervention (n = 247) | Control (n = 153) |
| --- | --- | --- |
| Age |  |  |
| 21-30 years | 42 (17.0) | 42 (27.5) |
| 31-40 years | 179 (72.5) | 106 (69.3) |
| >41 years | 26 (10.5) | 5 (3.2) |
| Italian nationality | 234 (94.7) | 139 (90.9) |
| Education |  |  |
| Primary school | 2 (0.8) | 0 (0.0) |
| Secondary school | 8 (3.2) | 2 (1.3) |
| High school | 70 (28.3) | 49 (32.0) |
| Academic degree | 167 (67.6) | 102 (66.7) |
| Health professionals | 55 (22.3) | 24 (15.7) |
| Other prenatal class participation | 49 (20) | 30 (19.6) |
| First pregnancy | 219 (89.4) | 130 (85.5) |
| Twin pregnancy | 4 (1.6) | 3 (2.0) |
| Scheduled C-section | 11 (4.5) | 8 (5.2) |

All data are reported as count (percentage).

**Table 2.** Baseline newborn characteristics.

|  | Intervention (n = 211) | Control (n = 128) |
| --- | --- | --- |
| Sex, F (%) | 113 (53.8) | 66 (51.2) |
| GA in weeks, mean ± SD | 39.6 ± 1.3 | 39.4 ± 1.5 |
| Weight at birth, g ± SD | 3300.8 ± 458.8 | 3250.4 g ± 427.9 |
| APGAR 1 min, mean ± SD | 9.0 ± 0.5 | 8.8 ± 1.1 |
| APGAR 5 min, mean ± SD | 9.8 ± 0.4 | 9.8 ± 0.4 |
| Delivery, n (%) |  |  |
| Vaginal | 177 (83.8) | 105 (82.1) |
| C-section | 11 (5.2) | 8 (6.2) |
| Emergency C-section | 23 (11.0) | 15 (11.7) |
| Labour duration, mean ± SD | 6.5 ± 6.2 | 7.2 ± 6.9 |
| Obstetric manoeuvres, n (%) | 16 (7.6) | 9 (7.0) |
| ODDI at birth (mean ± SD) | 101.6 ± 1.7 | 101.6 ± 1.7 |

GA: gestational age, ODDI: oblique diameter difference index, plagiocephaly: ODDI ≥ 104.

### Primary outcome - Craniometry

The ODDI score at the 3-month follow-up was evaluated using a cluster-adjusted ANCOVA model, controlling for baseline ODDI and hospital centres. There was no statistically significant difference in the follow-up ODDI score between the groups (Figure 3). The adjusted mean difference for the control group compared to the intervention group was −0.28 (95% CI: −0.73 to 0.17; p = 0.213). Furthermore, the linear mixed-effects model evaluating the clustering effect of the prenatal classes yielded an ICC of approximately zero (< 0.001), indicating that clustering did not significantly affect the variance of the primary outcome. A non-parametric sensitivity analysis using rank-transformed ODDI scores confirmed these findings (p = 0.169).

**Figure 3.**
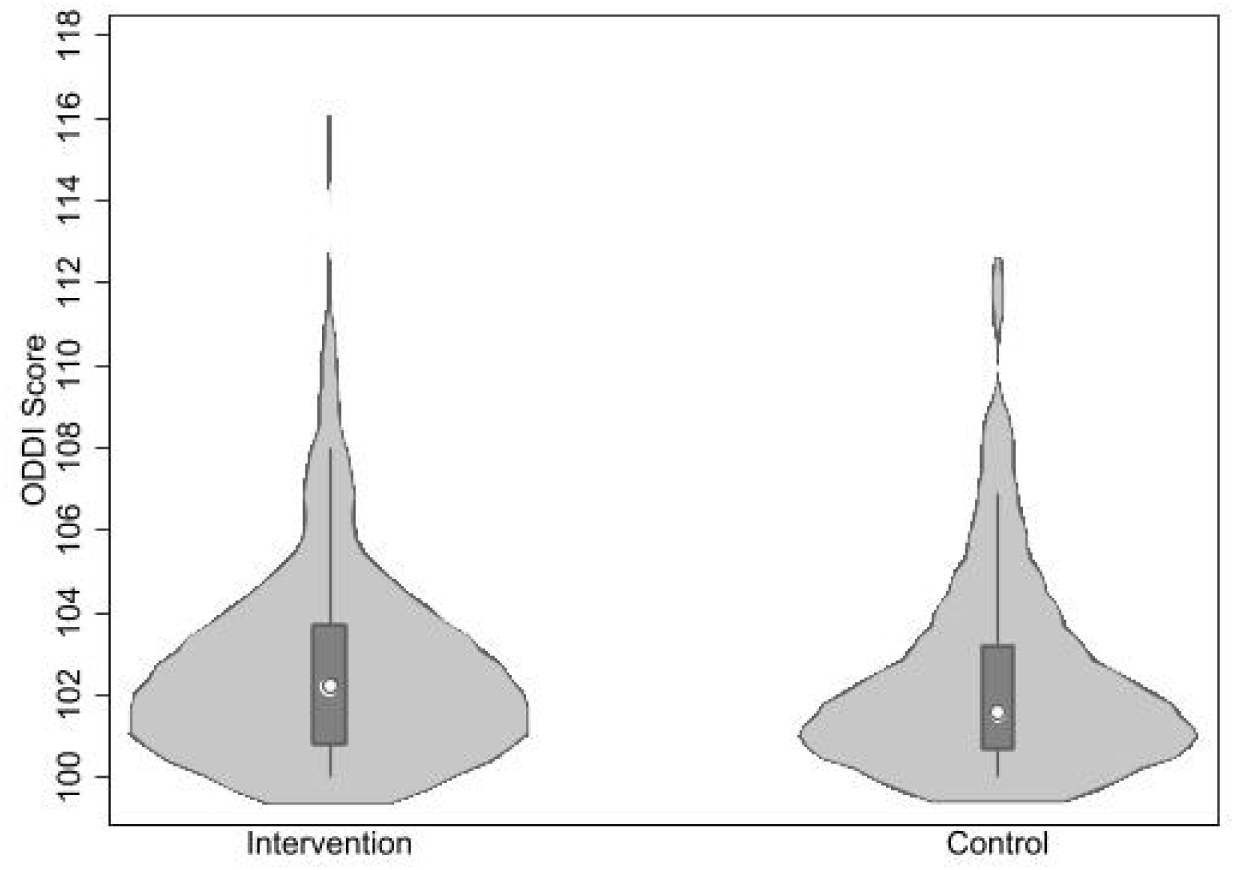
ODDI score at 3 months. The prevalence of plagiocephaly (defined as ODDI ≥ 104) at follow-up was 21.1% in the intervention group and 22.2% in the control group. The adjusted Poisson regression model revealed no significant difference in the risk of developing plagiocephaly between the control and intervention groups (adjusted Relative Risk [aRR] = 1.03; 95% CI: 0.70 to 1.51; p = 0.898).

### Secondary outcomes

There were no significant differences for secondary outcomes related to infant postural control and musculoskeletal assessments at 3 months (Figure 4), except for the spinal postural asymmetry, for which the infants in the control group had a 5.4% lower risk of exhibiting this posture compared to the intervention group (aRR = 0.95; 95% CI: 0.90 to 0.99; p = 0.033).

**Figure 4.**
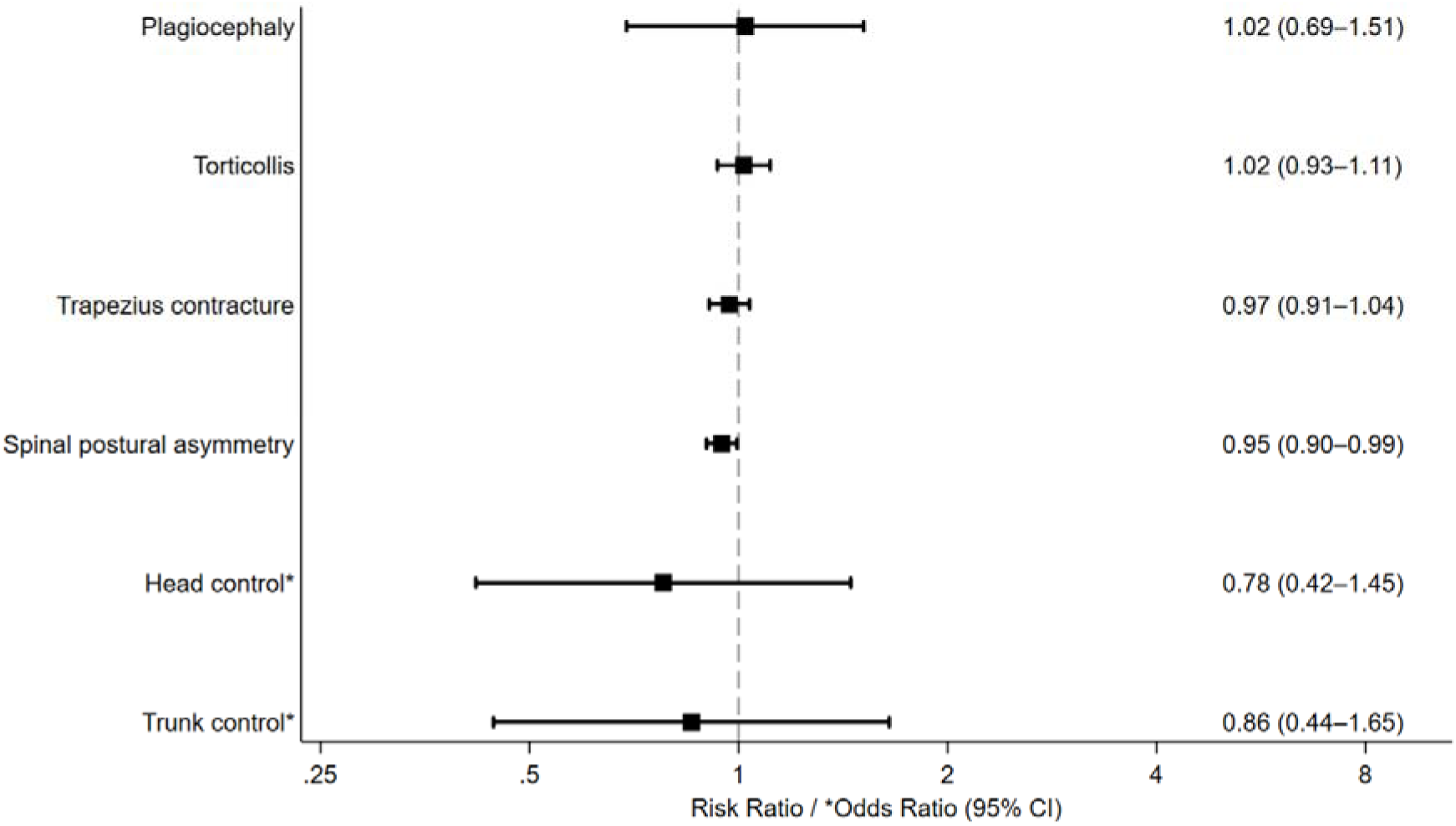
Secondary outcomes analyses. There were no significant differences in care practices between the two groups, either in position whilst at rest or in the use of equipment during transport.

### Perceived quality of the intervention

One hundred parents completed the questionnaire on the perceived quality of the intervention. The prenatal educational sessions were helpful, and the information provided was clear in 91% of respondents. 62% were already aware of the plagiocephaly condition. The greatest difficulties encountered in applying the learned strategies were managing positioning during sleep (31%) and tummy time (5%). 98% of respondents found the physical therapist’s presence during the prenatal class helpful, and 79% shared the information in the brochure with others.

## Discussion

This multicentre cluster RCT aimed to evaluate the effectiveness of a prenatal educational class on infant cranial asymmetry and posture-related disorders and is the first study on the prevention of POP conducted on the Italian paediatric population. The intervention did not result in a significant reduction in ODDI score or in the prevalence of POP at 3 months compared with standard care. Also, most secondary postural outcomes did not differ between groups, except for a slightly lower risk of a spinal postural asymmetry in the control group, the clinical relevance of which remains uncertain.

Because randomisation occurred during prenatal classes, a proportion of women assigned to the control group declined to provide consent for their newborns to be screened for eligibility at birth, potentially introducing selection imbalance. However, despite differences in consent rates between arms, baseline newborn characteristics were balanced, suggesting that the initial recruitment challenges in the control arm did not significantly affect internal validity.

A notable finding of this study was the low prevalence of POP (21.1% and 22.2%) across the entire cohort compared to current literature estimates, which range from 37.8%^14^ to 46.6%^21^ in similar age groups. Given that the questionnaire indicated prior awareness of the POP condition and that participants admitted to sharing experimental information with others, this may suggest contamination between the study arms. Moreover, given that the randomisation occurred within the same urban prenatal classes, mothers in the control group may have acquired intervention-related information also through direct social interactions or shared digital networks. Furthermore, the information-seeking behaviour prevalent among modern parents likely played a crucial role. Indeed, in the current technological landscape, parents are highly proactive in accessing online educational resources.^30–32^

Thus, it is plausible that mothers in the control group may have independently promoted the adoption of preventive behaviours (e.g., supervised prone positioning or active head repositioning). Furthermore, the families in the control group had agreed to take part in a study on the prevention of POP and may therefore have shown greater interest in the subject. Together, these factors may have reduced the measurable gap between the two groups and led to a ceiling effect in the educational intervention’s effectiveness.

An analysis of data on care practices implemented at home by the families involved in the study revealed no statistically significant differences between the two groups. This has likely contributed to the reduced prevalence of POP at 3 months, compared with data in the literature, thereby confirming the effectiveness of preventive measures for POP, but with no differences between the two groups, as this knowledge is likely shared between them.

Consistent results across various statistical methods strongly support our findings. Both the primary ANCOVA model and the non-parametric sensitivity analysis produced similar non-significant results, and the regression models indicated that the intervention did not significantly impact the clinical prevalence of plagiocephaly at 3 months. The high follow-up rate (97%) and the use of multiple imputation further reinforce our confidence in the accuracy of these results.

From a clinical and implementation perspective, these results suggest that a face-to-face prenatal educational session may offer limited incremental benefit in settings where baseline awareness and access to information are already high. Rather than replacing traditional educational models, these findings support a shift toward integrated strategies that combine prenatal guidance with postnatal reinforcement and targeted support for higher-risk dyads. In parallel, the role of physiotherapists may increasingly include curating and validating accessible, evidence-based information to ensure that widely available digital content translates into safe and effective caregiving practices.

## Conclusions

The preventive strategies adopted in both study groups show a reduction in the prevalence of POP compared to the average values reported in the scientific literature. The lack of superiority of the structured educational intervention may signal a shift in the landscape of parental education. Indeed, the resource-intensive face-to-face education model for low-risk infants may provide limited additional benefit in settings where parental awareness and access to educational resources are already high. Thus, we propose a paradigm shift in which clinicians move from being primary transmitters of basic knowledge to stewards of digital content quality, for example, by distributing the brochure to all families before they are discharged from the maternity unit and monitoring adherence to educational standards. By directing parents to validated, evidence-based online resources, healthcare systems can achieve similar preventive outcomes at significantly lower organisational costs.

## Data Availability

All data produced in the present study are available upon reasonable request to the authors

## Ethics approval

Meyer Children’s Hospital IRCCS Ethics Committee (24/12/2019).

## Patient involvement

Beyond the families who kindly participated in the study, a group of parents not included in the trial was involved in assessing the educational documents that were subsequently used in the trial.

## Acknowledgements

The authors thank Irene Frisano, the physiotherapist who explained the preventive measures in the educational video used in this trial, which is included in the appendix. We are also grateful to our colleagues, Valentina Di Vuolo, Claudia Artese, Marta Cattabiani, Cecilia Ortolani, Valentina Penazzi, Elena Ciavatta, Chiara Ricciotti, Giulia Anelli, Maria Teresa Camusso, Alice De Vincenzi, Fabiola Durante and Emma Cerioni for their contributions to this research.

## Declaration of generative AI and AI-assisted technologies in the manuscript preparation process

During the preparation of this work, the authors used Grammarly (Superhuman Platform Inc., 2023) to correct grammar, spelling, and punctuation, and to improve clarity. After using this tool, the authors reviewed and edited the content as needed and take full responsibility for the published article.

